# The Mexican dataset of a repetitive transcranial magnetic stimulation clinical trial on cocaine use disorder patients: SUDMEX TMS

**DOI:** 10.1101/2023.06.21.23291661

**Authors:** Diego Angeles-Valdez, Jalil Rasgado-Toledo, Viviana Villicaña, Alan Davalos-Guzman, Cristina Almanza, Alfonso Fajardo-Valdez, Ruth Alcala-Lozano, Eduardo A. Garza-Villarreal

## Abstract

Cocaine use disorder (CUD) is a worldwide problem with severe health and socio-economic consequences, which results in behavioral, cognitive, and neurobiological disturbances. Consensus on treatments are still under discussion, however, repetitive transcranial magnetic stimulation (rTMS) has been proposed as a promising treatment for medication-resistant disorders, including substance use disorders. Here, we describe the Mexican dataset of an rTMS clinical trial in patients with CUD (SUDMEX-TMS), a longitudinal dataset of 54 CUD participants (8 female) with five timepoints: baseline (T0), two weeks (T1), three months (T2), six months (T3) follow-up, and twelve months (T4) follow-up. Clinical rTMS treatment consisted of a double-blinded randomized clinical trial (n = 24 sham/30 active) for 2 weeks and open label afterwards, and includes demographic, clinical, and cognitive measures, as well as magnetic resonance imaging (MRI) acquisition in all timepoints: 1) structural (T1-weighted), 2) functional (resting state fMRI), and 3) multishell high-angular resolution diffusion-weighted (DWI-HARDI) sequences. The present dataset could be used to examine the impact of rTMS on CUD participants in clinical, cognitive, and multimodal MRI metrics in a longitudinal design.

## Background and Summary

Cocaine use disorder (CUD) is a worldwide public health problem with severe socio-economic consequences ^1^. Clinical outcomes include attention, learning, and working memory deficit, impulsivity, and brain alterations ^2, 3^. Thus, developing effective treatments has been a major concern in clinical research. Pharmacological approaches along with psychosocial therapy are currently the standard treatment with low to moderate efficacy ^4^. Repetitive transcranial magnetic stimulation (rTMS) has gained popularity as a novel therapeutic intervention to reduce cocaine use and craving symptoms ^5–7^.

Current research is focused on trying to affect circuits underlying different addiction-related processes such as cue reactivity ^8^. Repeated TMS can activate these circuits and seems to induce neuroplastic changes in the meso-cortico-limbic system that can alter activity and connectivity ^9^. In addition, the use of neuroimaging techniques such as magnetic resonance imaging (MRI) has helped to study these neurobiological outcomes in the search for biomarkers of disorder status and treatment outcome.

Our dataset stems from our placebo-controlled double-blind randomized clinical trial (RCT) using rTMS as an add-on to treatment as usual (TAU). The main advantage of our dataset is that we acquired longitudinal psychiatric interviews with standard clinical assessment and multimodal MRI sequences, including multi-shell diffusion-weighted imaging (see box 1). A participants checklist with the specific MRI sequences acquired can be found in supplementary material. To date, the present dataset has been used to examine the short and long-term clinical benefits of rTMS and their impact on functional connectivity ^10^, to identify cognitive deficits of CUD participants by machine learning algorithms ^11^, to improve diffusion MRI segmentation methods using deep learning ^12^, to predict clinical outcomes using microstructural changes ^13^, and to identify a generalizable functional connectivity signature characterizes brain dysfunction in cocaine use disorder ^14^ . Altogether, this dataset could serve for the study of the longitudinal impact of rTMS as a promising add-on treatment for CUD, as well as to test new neuroimaging algorithms and analysis techniques.

## Method

### Participants

From a sample of n = 117 patients,54 patients were included in the study. Reasons for the dropouts are in Supplementary Material. The study ran from May 2017 to September 2019. The study was conducted at the Clinical Research Division of the National Institute of Psychiatry in Mexico City, Mexico. The study was approved by the Ethics Committee of the Instituto Nacional de Psiquiatría “Ramón de la Fuente Muñiz” (CEI/C/070/2016) and is in accordance with the latest version of the Declaration of Helsinki. The trial was registered at ClinicalTrials.gov (NCT02986438). All participants provided oral and written informed consent. Cocaine dependence was diagnosed in CUD patients using the MINI International Neuropsychiatric Interview-Plus Spanish version 5.0.0 ^15^. Sample size was calculated using G*Power ^16^, for a 2 x 2 ANOVA (calculated from craving changes in previous cocaine rTMS studies ^17^), to attain 80% power at α = 0.05. Demographic characteristics between groups are summarized in Table 1.

**Table 1.** Demographic measures between groups.

|  | CUD Groups |  | Statistic |
| --- | --- | --- | --- |
|  | Sham (n = 23) | Active (n = 30) |  |
| **Age** |  |  |  |
| | 33.35 ± 8.15 | 36.07 ± 6.82 | $t(42.61) = -1.29, p = 0.20$ |
| **Sex** |  |  |  |
| Male | 20 (86.95%) | 25 (83.33%) | $\chi^2(1) = 7.13e-32, p = 1$ |
| Female | 3 (13.05%) | 5(16.67%) |  |
| **Education in years** |  |  |  |
| | 13.40 ± 2.83 | 12.90 ± 3.06 | $t(49.09) = 0.60, p = 0.54$ |
| **Monthly Income, MXN** |  |  |  |
| | 6591.30 ± 11386.63 | 5116.67 ± 6319.76 | $t(32.25) = 0.55, p = 0.58$ |
| **Main substance of use** |  |  |  |
| Crack cocaine | 20 (86.95%) | 29 (96.6%) | $\chi^2(1) = 0.64, p = 0.42$ |
| Cocaine | 3 (13.05%) | 1 (3.4%) |  |
| **Onset age of cocaine use** |  |  |  |
| | 22.52 ± 6.76 | 22.77 ± 5.8 | $t(43.38) = -0.13, p = 0.89$ |
| **Years of cocaine use** |  |  |  |
| | 9.59 ± 7.61 | 12.07 ± 7.68 | $t(47.70) = -1.17, p = 0.24$ |

Continuous variables are reported as mean ± SD, and nominal as number (percentage from group): two-sample t-test and $\chi^2$ was performed for each variable; no variables, n.a. : not applicable; CUD: cocaine use disorder.

### Experimental design

The study consisted of four stages: 1) screening interview confirmed the criteria for substance use disorder (SUD) by a trained psychiatrist; 2) participants underwent a full clinical evaluation and initial MRI acquisition (baseline or T0); 3) Acute stage (RCT) where patients underwent regularly scheduled sessions (Active or Sham rTMS) for 10 days over 2 weeks (T1). The patients who received sham were invited to receive the active treatment, that is 10 additional days in a period of two weeks (T1-4); 4) finally, the open-label maintenance phase three months (T2), six months (T3) and 12 months (T4). Clinical and MRI data were collected at the time of, at four weeks only sham patients (T1-4), The study design is detailed in Figure 1.

**Figure 1.**
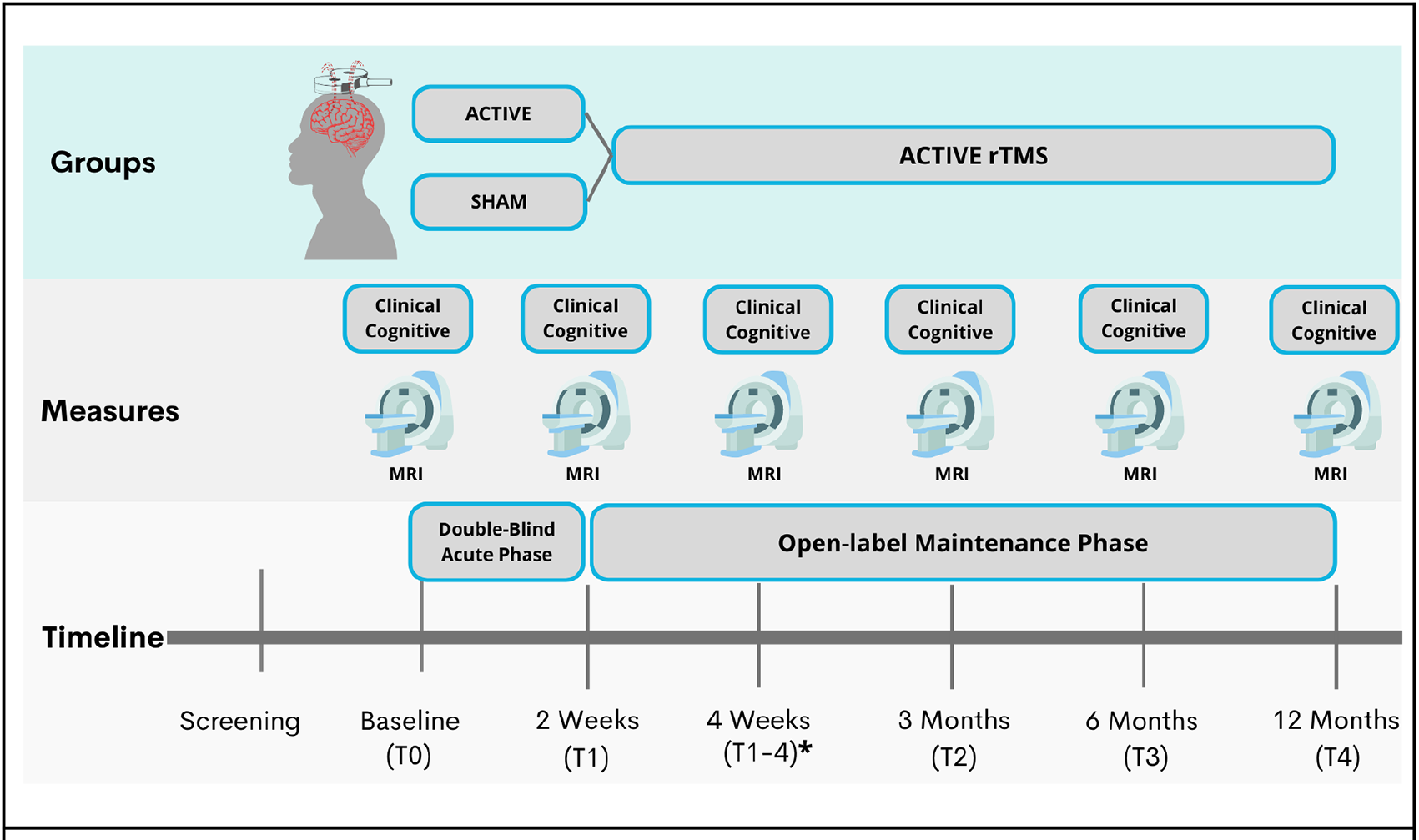
SUDMEX-TMS experimental design. Clinical and MRI data were collected at the time of baseline (T0), at two weeks (T1), three months (T2), six months (T3) and 12 months (T4); **\*** at four weeks only sham patients (T1-4).

### Study dropout

Of the 54 recruited patients, 30 were randomly allocated to active treatment and 24 to sham rTMS. Five patients assigned to active rTMS and four patients assigned to sham discontinued the study, leaving 24 patients in the Active group and 20 in the Sham group who completed the acute phase. Finally, in the double-blind phase, 14 patients in the Sham group opted for compassionate use and received 2 weeks of acute phase rTMS treatment after the sham. In the maintenance phase: 1) 20 patients (15 initially allocated to Active and 5 to Sham) finished 3 months of twice-weekly rTMS sessions (T2); 2) 15 patients (initially 10 Active and 5 Sham) finished 6 months of rTMS sessions (T3); and 3) 7 patients (initially 4 Active and 3 Sham) finished 12 months of twice-weekly rTMS sessions (T4). Due to substantial attrition at T1 (2 weeks), when the study was at ∼30% completion, we changed the maintenance phase to last 6 months instead of 12 months for new participants after approval by the ethics committee. None of the patients who interrupted the treatment at any time reported adverse effects of rTMS.

### Magnetic Resonance Imaging Acquisition

MRI sequences were acquired using a Philips Ingenia 3 T MR system (Philips Healthcare, Best, The Netherlands, and Boston, MA, USA), with a 32-channel dS Head coil. The order of the sequences was the following for the single session: 1) resting state (rs-fMRI), 2) T1-weighted (T1w), and 3) High Angular Resolution Diffusion Imaging (DWI-HARDI). This order was maintained across participants. Before the MRI acquisition, the amount of alcohol in participants’ blood was measured using a breathalyzer alcohol test and other substances were measured using breath alcohol test and *Instant-view^TM^* multi drug urine test. The total scan time was approximately 50 min. During the study, the participants were fitted with MRI-compatible headphones and goggles.

### Anatomical images

T1-weighted images were acquired using a 3D FFE SENSE sequence, TR/ TE = 7/3.5 ms, FOV = 240 mm^2^, matrix = 240 × 240 mm, 180 slices, gap = 0, plane = Sagittal, voxel = 1 × 1 × 1 mm (5 participants were acquired with a voxel size = 0.75 × 0.75 × 1 mm).

### Diffusion-weighted imaging

The DWI-HARDI was a spin echo (SE) sequence, with TR/TE =9000/127 ms, FOV=230 mm^2^ (for 4 participants=224mm^2^), matrix=96 x 96 (for 4 participants=112 x 112), number of slices=57 (for 4 participants=58), gap=0, plane=axial, voxel=2.4 x 2.4 x 2.5 mm (for 4 participants=2 x 2 x 2.3 mm), directions: 8 = b0, 32 = b-value 1,000 s/mm^2^ and 96 = b-value 2,500 s/mm^2^ (for 4 participants: 96 = b-value 3,000 s/mm^2^), with a total of 136 directions. We acquired a DWI-HARDI with an opposite direction for field mappings using a SE EPI sequence with the following parameters: TR/TE=9000/127ms, flip angle=90°, matrix=128 x 128 (for 4 participants=112 x 112), voxel size=1.8 x 1.8 x 2.5mm (for 4 participants=2 x 2 x 2.3mm), number of slices=57 (for 4 participants=58), phase encoding direction=PA. A total of 7 b0 volumes were acquired.

### Resting-state functional MRI

Resting state fMRI sequences were acquired using a gradient recalled (GE) echo planar imaging (EPI) sequence with the following parameters: dummies = 5, repetition time (TR)/echo time (TE)=2000/30.001ms, flip angle=75°, matrix=80x80, field of view=240mm^2^, voxel size=3 x 3 x 3mm, slice acquisition order=interleaved (ascending), number of slices=36, phase encoding direction=AP. The total scan time of the rs-fMRI session was 10 min, with a total of 300 volumes acquired. All participants were instructed to keep their eyes open, and to relax while not thinking about anything in particular. We used MRI-compatible goggles (fiber optic glasses SV-7021, Avotec) to show the participants a fixation cross (white cross with black background), and we used the included eye-tracking camera to prevent participants from falling asleep during this sequence. If the participants closed their eyes for more than 10 seconds, we would wake them up using the communication through the headphones, reminding them to try to not fall asleep for the 10 minutes the sequence lasted, and we re-started the sequence over. We acquired an opposite direction sequence for field mapping, using the same GE-EPI sequence with the following parameters: TR/TE=2000/30ms, flip angle=75°, matrix=80x80, voxel size=3x3x3mm, number of slices=36, phase encoding direction=PA. A total of 4 volumes were acquired.

### Transcranial magnetic stimulation

We performed a placebo-controlled double-blind randomized controlled trial (RCT) with parallel groups (Sham/Real) for 2 weeks (acute phase) and an open-label maintenance phase for 6 months. For the acute phase, we used a MagPro R301Option magnetic stimulator and a figure-of-eight B65-A/P coil (MagVenture, Alpharetta, GA); for the maintenance phase, we used a MagPro R30 stimulator and a figure-of-eight MCF-B70 coil (MagVenture). The acute phase comprised 10 weekdays of 5,000 pulses per day (two sessions of 50 trains at 5 Hz, 50 pulses/train, 10 s inter-train interval, and 15 min inter-session interval). The maintenance phase comprised 3 and 6 months of 5,000 pulses per day, 2 sessions per week. The maintenance phase comprised two 5-Hz excitatory frequency (5000 pulses per day) sessions per week. The stimulation was delivered at 100% motor threshold to the left Dorso-Lateral Prefrontal Cortex (lDLPFC). The motor threshold was determined in each patient as described by Rossini et al. ^18^. We used a vitamin E capsule as a fiducial during MRI acquisition to identify the stimulation cortical target due to a lack of a brain navigator. In all rTMS sessions, we used either the 5.5 cm anatomic Rule or the Beam F3 method. We changed to the superior Beam F3 method after the first 16 participants to improve lDLPFC localization ^19^. Electrodes were applied to all patient’s left temporalis muscles to simulate muscular contraction in the sham group, enhancing the sham and blinding, you can find more details in the supplementary material.

### Data Records

#### MRI Format Organization

The organization of the dataset follows the Brain Imaging Data Structure (BIDS, v. 1.0.1) (https://bids-specification.readthedocs.io/), commonly used to facilitate data sharing and project unifications by folder and file name structure according to sequence modality ^20^. Each MRI acquisition is shared in the Neuroimaging Informatics Technology Initiative (Nifti) format converted from Digital Imaging and Communication In Medicine (DICOM) using dcm2bids v.2.1.4. ^21^, along with data descriptions and metadata in JavaScript Object Notification files. The dataset is available and hosted on the OpenNeuro Data sharing platform (https://openneuro.org/datasets/ds003037) ^22^. Anonymization of the dataset was performed using pydeface to remove facial features ^23^.

#### Quality Control

To assess the quality of T1w and rsfMRI data, we used the MRI Quality Control tool (MRIQC) v.0.15, an automatic prediction of quality with visual reports of each sequence ^24^. For Diffusion-weighted images, we performed automated diffusion MRI QC (FSL EDDY_QC) to extract QC metrics sensitive and specific to artifacts ^25^.

#### Clinical measures

Patients were evaluated with a set of paper-based clinical tests at each session. The tests included were: 1) Mini International Neuropsychiatric Interview – Plus (MINI-Plus), 2) Addiction Severity Index (ASI), 3) Structured Clinical Interview for DSM-IV Axis II Personality Disorders (SCID-II), 4) Symptom Checklist-90-Revised (SCL-R), 5) Cocaine Craving Questionnaire General (CCQ-G) and Now (CCQ-N), 6) World Health Organization Disability Assessment Schedule 2.0 (WHODAS), 7) Barratt Impulsiveness Scale v. 11 (BIS-11), 8) Edinburgh Handedness Inventory Short Form, 9) Hamilton Depression Rating Scale (HDRS), 10) Hamilton Anxiety Rating Scale (HARS), 11) Pittsburgh Sleep Quality Index (PSQI), 12) Cocaine Craving visual analog scale (VAS). These clinical tests were carried out by trained mental health psychologists and psychiatrists, in a quiet room without distractions. Summary of the acquired data are summarized in Table 2.

**Table 2.** Summary of the acquired data for clinical measures, cognitive measures and MRI sequences.

| Acquisitions |  |  |
| --- | --- | --- |
| Clinical measures | Cognitive measures | MRI sequences |
| • Instant-view urine test<br>• MINI-Plus<br>• ASI<br>• SCID-II<br>• SCL-R Revised<br>• CCQ General & CCQ Now<br>• WHODAS<br>• BIS-11<br>• EHI short<br>• HDRS<br>• HARS<br>• PSQI<br>• VAS | • Berg's Card Sorting Test<br>• Flanker task<br>• Go/No-go task<br>• Letter number sequencing<br>• Digit span backward<br>• Iowa gambling task<br>• Tower of London<br>• Reading mind in the eyes | • rs-fMRI (10 min)<br>• Structural scan (T1-weighted)<br>• High Angular Resolution Diffusion Imaging (DWI) |

Mini International Neuropsychiatric Interview - Plus; MINI- Plus, Addiction Severity Index; ASI, Structured Clinical Interview for DSM-IV Axis II Personality Disorders; SCID-II, Symptom Checklist-90-Revised; SCL-R, Cocaine Craving Questionnaire General CCQ-G and Now CCQ-N, World Health Organization Disability Assessment Schedule 2.0; WHODAS, Barratt Impulsiveness Scale v. 11; BIS-11, Edinburgh Handedness Inventory Short Form; EHI short, 9) Hamilton Depression Rating Scale (HDRS), 10) Hamilton Anxiety Rating Scale (HARS), 11) Pittsburgh Sleep Quality Index (PSQI), 12) Cocaine Craving visual analogue scale (VAS).

#### Mini International Neuropsychiatric Interview – Plus (M.I.N.I.-Plus)

The M.I.N.I. is a comprehensive diagnostic psychiatric interview designed to assess the 17 most common psychiatric disorders of the Axis I in DSM-IV and the ICD-10 ^26^. We used the official Spanish translation version 5.0. of the M.I.N.I.-Plus, an extended version that includes a total of 23 psychiatric disorders. The interview consists of a set of structured questions that require a yes or no answer. These questions are designed to cover all the diagnostic criteria of the different psychiatric disorders. The interview is divided into modules that correspond to the diagnostic category which was applied by qualified psychiatrists.

#### Addiction severity index (ASI)

The ASI is a semi-structured interview designed to assess the lifetime and the past-30-days status in several functional domains: alcohol and drug use, medical and psychiatric health, employment, self-support, family relations, and illegal activities ^27^. We used the Spanish translation of the revised 5th edition ^28^.

#### Structured clinical interview for dsm-iv axis ii personality disorders (SCID-II)

The SCID-II is a psychiatric interview that allows the diagnosis of DSM-IV personality disorders ^29^. The instrument is a self-administered questionnaire that asks the DSM-IV Axis-II criteria in the form of yes/no questions. We only applied for this SCID-II questionnaire. First, it briefly characterizes the typical behavior, relationships, and capacity for self-reflection of the interviewee and then assesses each of the personality disorders. We used the official Spanish translation ^30^.

#### Symptom checklist-90-revised (SCL-90-R)

The SCL-90-R is a self-report measure of psychological symptoms and distress ^31^. It includes 9 symptom dimensions (somatization, obsessive-compulsive, interpersonal sensitivity, depression, anxiety, hostility, phobic anxiety, paranoid ideation, and psychoticism) and 3 global indices of distress: Global Severity Index (GSI), Positive Symptom Distress Index (PSDI) and Positive Symptoms Total (PST). To score the instrument, an average of all the items for each symptom dimension is obtained. The GSI is calculated by calculating a global average of 90 items. The PSDI is the amount of non-zero responses given by the participant, while PST is calculated by the sum of the 90 items’ scores divided by the PSDI. In general, for the 9 symptom dimensions and the 3 global indices of distress, values above the 90th percentile are considered high, indicating a person at risk. We used a Spanish translation ^32^.

#### Cocaine craving questionnaire general (CCQ-General) and Now (CCQ-Now)

The CCQ is a 45-item questionnaire that explores cocaine craving among patients ^33^. The CCQ-General asks participants to rate their cravings over the previous week. The CCQ-Now asks participants about their cravings at the moment of assessment. Both versions of the CCQ consist of the same items, written in different tenses. The items are related to the following contents: the desire to use cocaine, intention and planning to use cocaine, anticipation of a positive outcome, the anticipation of relief from withdrawal or dysphoria, and lack of control overuse. We used the CCQ-General Spanish translation that was validated in the Mexican population ^34^.

#### World health organization disability assessment schedule 2.0 (WHODAS 2.0)

The WHODAS is an instrument designed to evaluate functional impairments or disabilities of patients with psychiatric disorders. The WHODAS 2.0 is an updated version of instrument ^35^. It provides information on four areas: Personal Care, Occupation, Family/Housing and Social Functioning. We used the official 36-item, self-administered, Spanish translation of this version ^35, 36^.

#### Barratt impulsiveness scale version 11 (BIS-11)

The 11th version of the Barratt Impulsivity Scale is one of the most widely used instruments for assessing impulsivity ^37^. This version of the scale consists of 30 items that describe impulsive and non-impulsive behaviors related to 3 main categories: attentional, motor, and non-planning/impulsiveness. We interpret the scores based on the following classification: highly impulsive (total score ≥ 72), within normal limits for impulsiveness (total score 52–71), extremely over-controlled, or has not honestly completed the questionnaire (total score <52) ^38^. We used the Spanish translation of the BIS-11 ^39^.

#### Hamilton Depression Rating Scale (HDRS)

The Hamilton Rating Scale for Depression was used to provide a measure of the severity of depression. The version we used is one of 17 items. Its content focuses on the behavior of depression, with vegetative, cognitive, and anxiety symptoms having the greatest weight in the total calculation of the scale. The cutoff points to define severity are no depression (0-7); mild depression (8-16); moderate depression (17-23); and severe depression (≥24) ^40^.

#### Hamilton Anxiety Rating Scale (HARS)

This scale assesses the severity of anxiety globally and is useful for monitoring response to treatment. It consists of 14 items, to measure 13 anxious signs and symptoms, and the last one that evaluates the patient’s behavior during the interview. The interviewer scores from 0 to 4 points for each item, assessing both its intensity and frequency. The total score is the sum of those of each of the items. The range is from 0 to 56 points. The optimal HAM-A score ranges were: no/minimal anxiety ≤ 7; mild anxiety = 8-14; moderate = 15-23; severe ≥ 24 ^41^ .

#### Pittsburgh Sleep Quality Index (PSQI)

This instrument has been developed to measure sleep quality in patients with psychiatric disorders. It is made up of 24 items, although only 19 are taken into account for its correction. In addition, it is divided into 7 dimensions: Subjective sleep quality, Sleep latency. Duration of sleep, Usual sleep efficiency, Sleep disturbances, Use of medication, Daytime dysfunction. It is answered with a Likert-type scale that goes from 0 to 4. For its correction, a sleep profile is obtained in each of the dimensions ranging from 0 to 3 and a total score that can range from 0 to 21 ^42^.

#### Cocaine Craving visual analogue scale (VAS)

It is an instrument for the subjective assessment of the participant’s current craving. The visual scale consists of a continuous line of 10 cm (including 2 decimal places), where the left end refers to *“no craving”* and the right end to *“the most intense craving”* and the participants must mark with a cross the intensity of his craving he currently feels ^43^.

#### Instant-view^TM^ multi-drug urine test

Performed to identify the possible presence of substances of abuse in participants prior to performing the MRI study. This test was performed with an Instant-view^TM^ Multi Drug Urine Test (catalog number 03-3635) from Alpha Scientific Designs Inc (California, USA) https://www.alfascientific.com/products/multi-drug-urine-tests/, using the lateral flow chromatographic immunoassay technique. The substances detected and their cut-off points are as follows: Amphetamines (1000 ng/mL), Benzodiazepines (300 ng/mL), Cocaine (300 ng/mL), Methamphetamine (1000 ng/mL), Morphine/Opiates (2000 ng/mL), Marijuana/Hashish (50 ng/mL).

#### Cognitive tests

Patients were administered a comprehensive neuropsychological assessment at the three stages in two different modalities: computer-based using the Psychology Experiment Building Language (PEBL) version 2.0 with Spanish translation ^44^ and paper-based: 1) Berg’s Card Sorting Test, 2) Flanker Task, 3) Go/No-Go task, and 4) Letter-number test ^44^, 5) Digit span backward, 6) Iowa gambling task, 7) Tower of London, and 8) Reading the Mind in the Eyes Test. Both the Digit-Span Backward and 9) Letter-Numbers Sequencing exams were administered on paper. The cognitive tests were applied by a licensed psychologist in a quiet setting to minimize distractions. The assessment was conducted after the MRI scan ^44^, with a total duration of 45 minutes per participant. The tests were the following:

#### Cognitive flexibility

BCST or Berg’s Card Sorting Test: The Wisconsin Card Sorting Test for computers consists of a deck of 128 cards. The participants are shown one card at a time to match them into categories displayed on the screen (shape, color, or amount). The goal is for the participants to figure out the rule for categorizing. The rule changes when ten appropriately matched cards have been found ^12, 13^.

#### Inhibition

The flanker task assesses response inhibition processes by identifying stimuli over the noise. In a target-distracting sequence, participants are required to react to the middle arrow in a rapidly changing row of arrows while ignoring the others ^45, 46^ Go/No-go task: In this task, participants must give a prompt answer to a go signal and hold back their response to a no-go signal. Reaction time (RT) and the capacity to block the response under no-go signals are measured ^47, 48^.

#### Working memory

Letter-number sequencing: Participants are told a set of letters and numbers, which they must recall and repeat in the correct order, beginning with the numbers and going alphabetically up from there ^49^.

Digit span backward: Participants must recite a list of numbers in the reverse order that has been supplied by the examiner. The level of difficulty rises every two items and ends after two consecutive unsuccessful answers ^49^.

#### Decision Making

Iowa gambling task (IGT): Participants look at four digital card decks. They need to turn over one at a time to complete 100 trials. They need to identify the pattern of beneficial or unfavorable outcomes (receiving or losing a specific amount of money) and make their choice. The goal is to maximize earnings, out of the initial given amount of $2,000 ^50, 51^.

Tower of London (ToLo) planning: This activity is frequently used to gauge planning ability and other executive functions. Participants must preplan a series of moves, one token at a time, to replicate the target model shown on the top board with the fewest number of movements ^52, 53^.

#### Theory of mind

The Reading the Mind in the Eyes Test (RMET): Participants are shown a grayscale image of the upper half face. They are required to choose from four options on the screen, which words describe the emotion or mental state of the image. A dictionary is available on the top screen in case participants are not familiar with any of the words ^54^

#### Technical Validation of the MRI

The automatic quality control of sMRI and rsMRI sequences was assessed using the MRIQC v.0.15 tool ^24^. For sMRI the extracted values were Signal-to-Noise Ratio (SNR) and Contrast-to-Noise Ratio (CNR) (Figure 2). SNR is related to the ratio of the mean voxel intensity of an image in contrast with the random noise intensity ^55^, whereas CNR measure is an extension of SNR that is not influenced by contrast changes ^56^.

**Figure 2.**
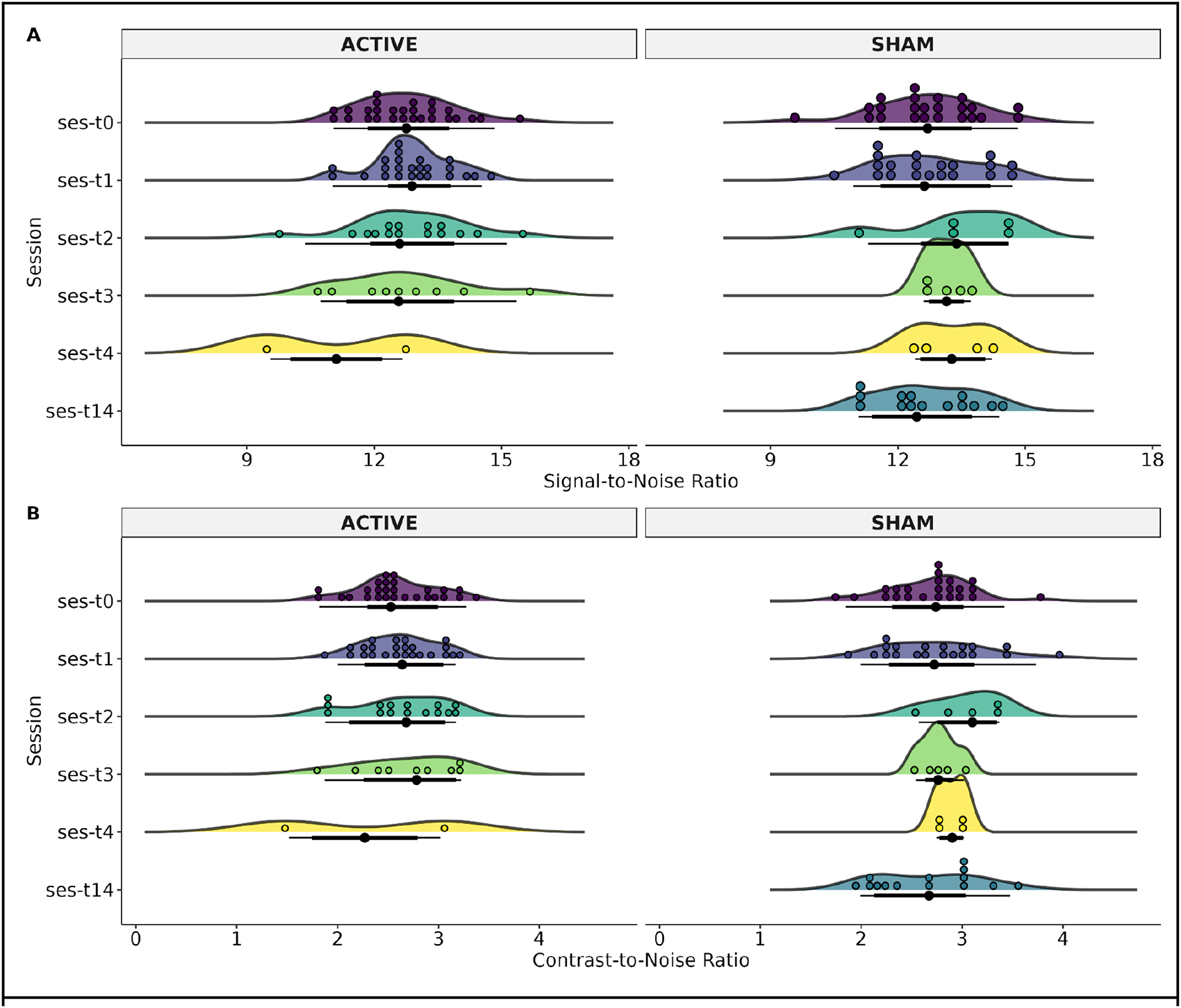
Status of the structural weighted image of each MRI-Session (ses): baseline (T0), at two weeks (T1), three months (T2), and six months (T3), and patients that had 12 months (T4). The T14 time was for patients in the Sham group who decided to continue the clinical trial with open-label rTMS. The T14 refers to 2 weeks after T1 (4 weeks after T0). A) Signal-to-Noise Ratio (SNR), and B) Contrast-to-Noise Ratio (CNR).

For rsMRI the extracted values were Framewise Displacement (FD) and temporal Signal-to-Noise Ratio (tSNR) (Figure 3). FD is the sum of parameters of translational and rotational realignment of head motion ^56, 57^, while tSNR is calculated as the division of the mean of the time series by its standard deviation, when spatial resolution increases, TSNR decreases ^58^.

**Figure 3.**
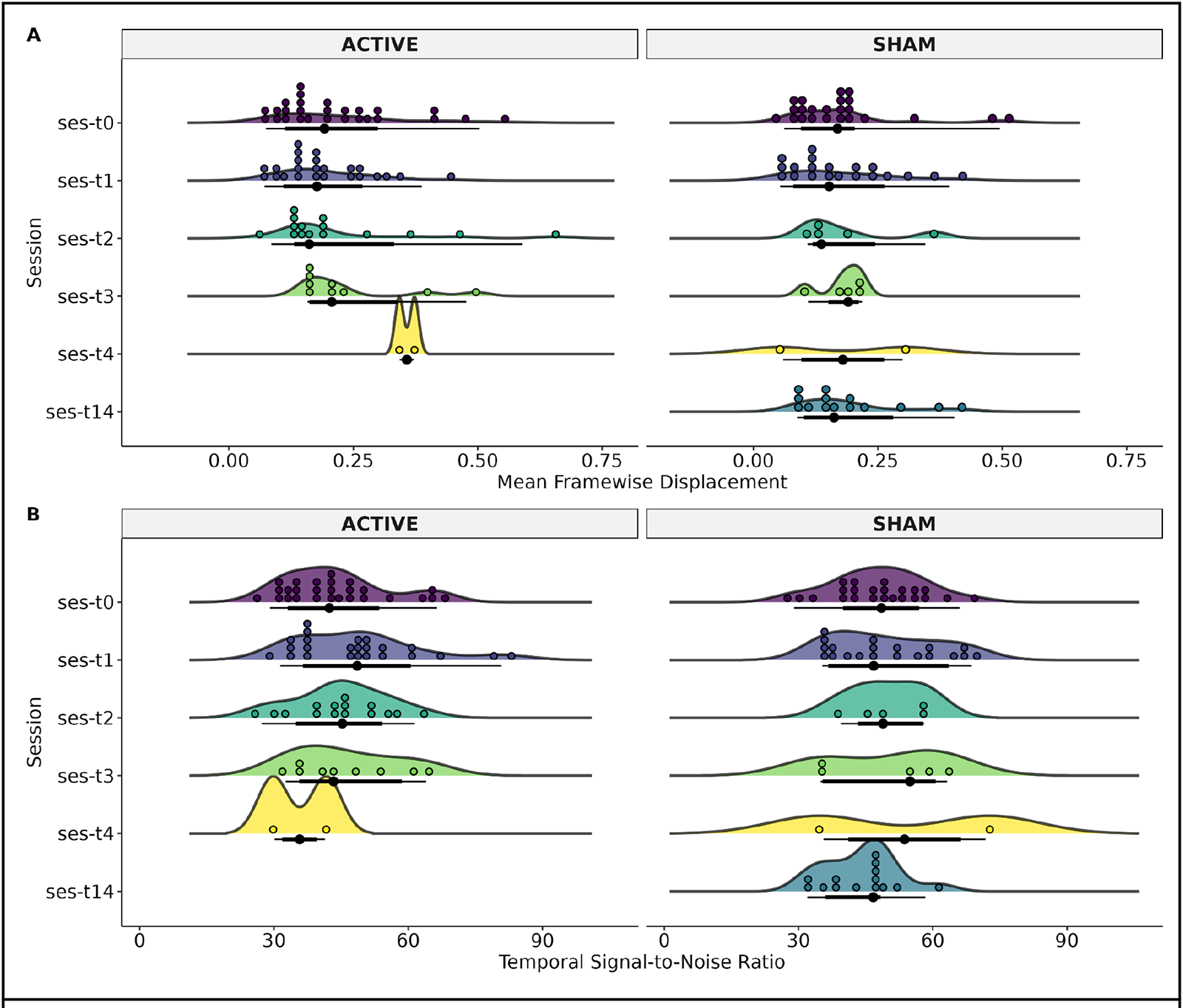
Status of the resting-state image of each MRI-Session (ses): baseline (T0), at two weeks (T1), three months (T2), and six months (T3), and patients that had 12 months (T4). The T14 time was for patients in the Sham group who decided to continue the clinical trial with open-label rTMS. The T14 refers to 2 weeks after T1 (4 weeks after T0). A) Mean Framewise Displacement (FD) and, B) Temporal Signal-to-Noise Ratio (tSNR).

For Diffusion-weighted images (dMRI), the quality of sequences was assessed using EDDY_QC a FSL tool ^25^, at a single level. The extracted values were related to motion: absolute (motion referenced to the middle time-point) and relative (motion compared with the previous time-point). The automated QC tool relies on EDDY ^59^, used to calculate the motion of dMRI data through volume-to-volume motion based on 6 parameters of the degree of freedom (Figure 4).

**Figure 4.**
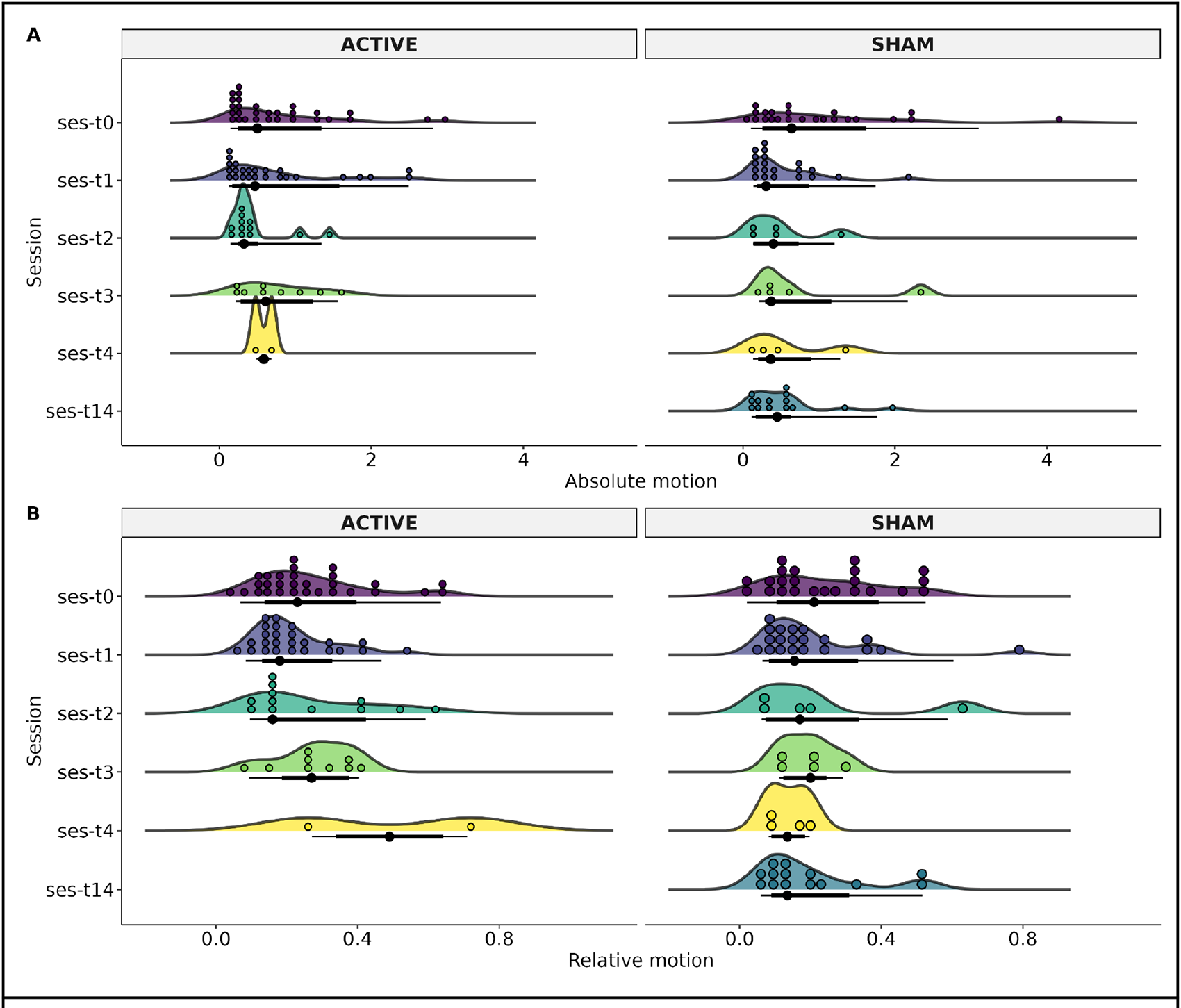
Participant motion of the diffusion-weighted image of each MRI-Session (ses): baseline (T0), at two weeks (T1), three months (T2), and six months (T3), and patients that had 12 months (T4). The T14 time was for patients in the Sham group who decided to continue the clinical trial with open-label rTMS. The T14 refers to 2 weeks after T1 (4 weeks after T0). A) Absolute motion, B) Relative motion.

#### Usage Notes

The present dataset consists of cocaine use disorder patients treated using rTMS therapy followed up to 12 months and divided into a sham group and an active rTMS group for the first 2 weeks. Uses include scientific/clinical research and academic purposes. Studies can be focused on the impact of rTMS over the active group in contrast to sham, on multimodal MRI data and/or clinical/cognitive measures. To use the present dataset in conjunction with larger-scale projects, such as those of the ENIGMA consortium, we recommend the use of an MRI-site harmonization technique such as ComBatHarmonization ^60^. Here, we provided QC information in order to facilitate data usage of which participants could be dismissed from data analysis. We recommend the use of artifact correction methods for preprocessing data due to the high motion of some subjects during scanning.

## Data Availability

All data produced in the present study are available upon reasonable request to the authors, and all data produced are available online at

https://zenodo.org/record/7126853

## Code Availability

The MRI dataset can be found in https://openneuro.org/datasets/ds003037/. Please download the latest available version as there may be updates. Clinical and cognitive measures are available in Zenodo https://doi.org/10.5281/zenodo.7126853.

## Acknowledgments

We thank the people who helped this project in one way or another: Ernesto Reyes-Zamorano, Francisco J. Pellicer Graham, Erik Morelos-Santana, Sarael Alcauter, Luis Concha, Bernd Foerster, and Alejandra López-Castro. We also thank Rocio Estrada Ordoñez and Isabel Lizarindari Espinosa Luna at the Unidad de Atención Toxicológica Xochimilco for all their help and effort. Finally, we thank the study participants for their cooperation and patience. This study was supported by public funds CONACYT FOSISS No. 0260971 and CONACYT No. 253072. Diego Angeles-Valdez is a doctoral student from Programa de Maestría y Doctorado en Psicología, Universidad Nacional Autónoma de México (UNAM) and received fellowship No. 1003596 from Consejo Nacional de Ciencia y Tecnología (CONACYT). Jalil Rasgado Toledo is a doctoral student from the Programa de Doctorado en Ciencias Biomédicas, Universidad Nacional Autónoma de México (UNAM) and received fellowship 858667 from CONACYT. This work received support from Luis Aguilar, Alejandro De León, and Jair García of the Laboratorio Nacional de Visualización Científica Avanzada, and also we appreciate the technical support by Leopoldo González-Santos

## Author contributions

E.G.V. originated the concept of the dataset, designed the study, provided funding and acquired pilots. R.A.L, D.A.V., and V.V. recruited patients. D.A.V. and V.V. acquired the MRI data. D.A.V., and V.V. implemented the cognitive tests. R.A.L. implemented and managed the clinical part of the study. D.A.V., J.R.T., and C.A., performed the analysis and technical validation. D.A.V., J.R.T., A.F.V. and D.A.V., J.R.T., and E.G.V. created, reviewed and approved the manuscript.

## Competing interests

The authors declare no conflict of interest.

## Notes

### Competing Interest Statement

The authors have declared no competing interest.

### Clinical Trial

NCT02986438

### Author Declarations

Ethics Committee of the Instituto Nacional de Psiquiatria - Ramon de la Fuente Muniz (CEI/C/070/2016)

## References

1. Peacock, A. et al. Global statistics on alcohol, tobacco and illicit drug use: 2017 status report. Addiction 113, 1905–1926 (2018).

2. Potvin, S., Stavro, K., Rizkallah, E. & Pelletier, J. Cocaine and cognition: a systematic quantitative review. J. Addict. Med. 8, 368–376 (2014).

3. Fryer, R. G., Jr, Heaton, P. S., Levitt, S. D. & Murphy, K. M. Measuring crack cocaine and its impact. Econ. Inq. 51, 1651–1681 (2013).

4. Kampman, K. M. The treatment of cocaine use disorder. Sci Adv 5, eaax1532 (2019).

5. Terraneo, A. et al. Transcranial magnetic stimulation of dorsolateral prefrontal cortex reduces cocaine use: A pilot study. Eur. Neuropsychopharmacol. 26, 37–44 (2016).

6. Politi, E., Fauci, E., Santoro, A. & Smeraldi, E. Daily sessions of transcranial magnetic stimulation to the left prefrontal cortex gradually reduce cocaine craving. Am. J. Addict. 17, 345–346 (2008).

7. Protasio, M. I. B. et al. The Effects of Repetitive Transcranial Magnetic Stimulation in Reducing Cocaine Craving and Use. *Addictive* Disorders & Their Treatment vol. 18 212–222 Preprint at https://doi.org/10.1097/adt.0000000000000169 (2019).

8. Steele, V. R. & Maxwell, A. M. Treating cocaine and opioid use disorder with transcranial magnetic stimulation: A path forward. Pharmacol. Biochem. Behav. 209, 173240 (2021).

9. Moretti, J., Poh, E. Z. & Rodger, J. rTMS-Induced Changes in Glutamatergic and Dopaminergic Systems: Relevance to Cocaine and Methamphetamine Use Disorders. Frontiers in Neuroscience vol. 14 Preprint at https://doi.org/10.3389/fnins.2020.00137 (2020).

10. Garza-Villarreal, E. A. et al. Clinical and Functional Connectivity Outcomes of 5-Hz Repetitive Transcranial Magnetic Stimulation as an Add-on Treatment in Cocaine Use Disorder: A Double-Blind Randomized Controlled Trial. Biological Psychiatry: Cognitive Neuroscience and Neuroimaging vol. 6 745–757 Preprint at https://doi.org/10.1016/j.bpsc.2021.01.003 (2021).

11. Jiménez, S. et al. Identifying cognitive deficits in cocaine dependence using standard tests and machine learning. Prog. Neuropsychopharmacol. Biol. Psychiatry 95, 109709 (2019).

12. Zhang, F. et al. Deep learning based segmentation of brain tissue from diffusion MRI. Neuroimage 233, 117934 (2021).

13. Rasgado-Toledo, J., Issa-Garcia, V., Alcalá-Lozano, R., Garza-Villarreal, E. A. & González-Escamilla, G. Cortical and subcortical connections change after repetitive transcranial magnetic stimulation therapy in cocaine use disorder and predict clinical outcome. Preprint at https://doi.org/10.1101/2022.09.29.22280253.

14. Zhao, K. et al. A generalizable functional connectivity signature characterizes brain dysfunction and links to rTMS treatment response in cocaine use disorder. medRxiv (2023) doi:10.1101/2023.04.21.23288948.

15. Ferrando, L., Bobes, J., Gibert, M., Soto, M. & Soto, O. M.I.N.I. PLUS Mini International Neuropsychiatric Interview. Versión en Español 5.0.0. Preprint at (1998).

16. Faul, F., Erdfelder, E., Buchner, A. & Lang, A.-G. Statistical power analyses using G*Power 3.1: tests for correlation and regression analyses. Behav. Res. Methods 41, 1149–1160 (2009).

17. Bolloni, C., Badas, P., Corona, G. & Diana, M. Transcranial magnetic stimulation for the treatment of cocaine addiction: evidence to date. Subst. Abuse Rehabil. 9, 11–21 (2018).

18. Rossini, P. M. et al. Non-invasive electrical and magnetic stimulation of the brain, spinal cord and roots: basic principles and procedures for routine clinical application. Report of an IFCN committee. Electroencephalogr. Clin. Neurophysiol. 91, 79–92 (1994).

19. Trapp, N. T. et al. Reliability of targeting methods in TMS for depression: Beam F3 vs.5.5 cm. Brain Stimul. 13, 578–581 (2020).

20. Gorgolewski, K. J. et al. The brain imaging data structure, a format for organizing and describing outputs of neuroimaging experiments. Sci Data 3, 160044 (2016).

21. Li, X., Morgan, P. S., Ashburner, J., Smith, J. & Rorden, C. The first step for neuroimaging data analysis: DICOM to NIfTI conversion. J. Neurosci. Methods 264, 47–56 (2016).

22. Markiewicz, C. J. et al. The OpenNeuro resource for sharing of neuroscience data. Elife 10, (2021).

23. Gulban, Nielson, Poldrack & Lee. poldracklab/pydeface: v2. 0.0. *Zenodo* https://doi.org.

24. Esteban, O. et al. MRIQC: Advancing the automatic prediction of image quality in MRI from unseen sites. PLoS One 12, e0184661 (2017).

25. Bastiani, M. et al. Automated quality control for within and between studies diffusion MRI data using a non-parametric framework for movement and distortion correction. Neuroimage 184, 801–812 (2019).

26. Sheehan, D. V. et al. The Mini-International Neuropsychiatric Interview (M.I.N.I.): the development and validation of a structured diagnostic psychiatric interview for DSM-IV and ICD-10. J. Clin. Psychiatry 59 Suppl 20, (1998).

27. McLellan, A. T., Luborsky, L., Woody, G. E. & O’Brien, C. P. An improved diagnostic evaluation instrument for substance abuse patients. The Addiction Severity Index. J. Nerv. Ment. Dis. 168, (1980).

28. McLellan, A. T. et al. The Fifth Edition of the Addiction Severity Index. J. Subst. Abuse Treat. 9, (1992).

29. First, M. B. User’s Guide for the Structured Clinical Interview for DSM-IV Axis II Personality Disorders: SCID-II. (American Psychiatric Pub, 1997).

30. First, M. B., Gibbon, M., Spitzer, R. L., Williams, J. B. W. & Benjamin, L. S. Entrevista Clínica Estructurada para los Trastornos de Personalidad del Eje II del DSM-IV SCID-II. Masson (1997).

31. Derogatis, L. R. The Symptom Checklist-90-R (SCL-90-R). Clinical Psychometrics Research (1975).

32. Casullo, M. M. & Castro Solano, A. Evaluación del bienestar psicológico en estudiantes adolescentes argentinos. *Revista de Psicología* vol. 18 35–68 Preprint at https://doi.org/10.18800/psico.200001.002 (2000).

33. Tiffany, S. T., Singleton, E., Haertzen, C. A. & Henningfield, J. E. The development of a cocaine craving questionnaire. Drug Alcohol Depend. 34, (1993).

34. Marín-Navarrete, R. et al. Validation of a cocaine craving questionnaire (CCQ-G) in Mexican population. Salud Ment. 34, 491–496 (2011).

35. World Health Organization. Measuring health and disability: manual for WHO Disability Assessment Schedule (WHODAS 2.0). (Word Health Organization, 2010).

36. World Health Organization. Medición de la salud y la discapacidad: manual para el cuestionario de evaluación de la discapacidad de la OMS: WHODAS 2.0. (World Health Organization / Servicio Nacional de Rehabilitación, 2015).

37. Patton, J. H., Stanford, M. S. & Barratt, E. S. Factor structure of the Barratt impulsiveness scale. J. Clin. Psychol. 51, (1995).

38. Fifty years of the Barratt Impulsiveness Scale: An update and review. Pers. Individ. Dif. 47, 385–395 (2009).

39. Oquendo, M. A. et al. Spanish adaptation of the Barratt Impulsiveness Scale (BIS-11). Eur. Psychiatry 15, 147–155 (2001).

40. Ramos-Brieva, J. A. & Cordero Villafáfila, A. [Relation between the validity and reliability of the Castillian version of the Hamilton Rating Scale for Depression]. Actas Luso. Esp. Neurol. Psiquiatr. Cienc. Afines 14, 335–338 (1986).

41. Lobo, A. et al. Validación de las versiones en español de la Montgomery-Asberg Depression Rating Scale y la Hamilton Anxiety Rating Scale para la evaluación de la depresión y de la ansiedad. Medicina Clínica vol. 118 493–499 Preprint at https://doi.org/10.1016/s0025-7753(02)72429-9 (2002).

42. Buysse, D. J., Reynolds, C. F., 3rd, Monk, T. H., Berman, S. R. & Kupfer, D. J. The Pittsburgh Sleep Quality Index: a new instrument for psychiatric practice and research. Psychiatry Res. 28, 193–213 (1989).

43. Nicholson, A. N. Visual analogue scales and drug effects in man. Br. J. Clin. Pharmacol. 6, 3–4 (1978).

44. Mueller, S. T. & Piper, B. J. The Psychology Experiment Building Language (PEBL) and PEBL Test Battery. *Journal of Neuroscience Methods* vol. 222 250–259 Preprint at https://doi.org/10.1016/j.jneumeth.2013.10.024 (2014).

45. Eriksen, B. A. & Eriksen, C. W. Effects of noise letters upon the identification of a target letter in a nonsearch task. Perception & Psychophysics vol. 16 143–149 Preprint at https://doi.org/10.3758/bf03203267 (1974).

46. Sanders, A. F. & Lamers, J. M. The Eriksen flanker effect revisited. Acta Psychol. 109, 41–56 (2002).

47. Gomez, P., Ratcliff, R. & Perea, M. A model of the go/no-go task. *Journal of Experimental Psychology: General* vol. 136 389–413 Preprint at https://doi.org/10.1037/0096-3445.136.3.389 (2007).

48. Wright, L., Lipszyc, J., Dupuis, A., Thayapararajah, S. W. & Schachar, R. Response inhibition and psychopathology: a meta-analysis of go/no-go task performance. J. Abnorm. Psychol. 123, (2014).

49. Wechsler, D. Wechsler Adult Intelligence Scale (WAIS-IV). (Psychological Corp, 2008).

50. Bechara, A., Damasio, A. R., Damasio, H. & Anderson, S. W. Insensitivity to future consequences following damage to human prefrontal cortex. Cognition 50, (1994).

51. Buelow, M. T. & Suhr, J. A. Construct validity of the Iowa Gambling Task. Neuropsychol. Rev. 19, (2009).

52. Krikorian, R., Bartok, J. & Gay, N. Tower of London procedure: a standard method and developmental data. J. Clin. Exp. Neuropsychol. 16, (1994).

53. Unterrainer, J. M. et al. Planning abilities and the Tower of London: is this task measuring a discrete cognitive function? J. Clin. Exp. Neuropsychol. 26, (2004).

54. Baron-Cohen, S., Wheelwright, S., Hill, J., Raste, Y. & Plumb, I. The ‘Reading the Mind in the Eyes’ Test revised version: a study with normal adults, and adults with Asperger syndrome or high-functioning autism. J. Child Psychol. Psychiatry 42, (2001).

55. McRobbie, D. W., Moore, E. A., Graves, M. J. & Prince, M. R. MRI from Picture to Proton. (Cambridge University Press, 2017).

56. Magnotta, V. A., Friedman, L. & FIRST Birn. Measurement of Signal-to-Noise and Contrast-to-Noise in the fBIRN Multicenter Imaging Study. J. Digit. Imaging 19, 140–147 (2006).

57. Power, J. D., Barnes, K. A., Snyder, A. Z., Schlaggar, B. L. & Petersen, S. E. Spurious but systematic correlations in functional connectivity MRI networks arise from subject motion. Neuroimage 59, 2142–2154 (2012).

58. Murphy, K., Bodurka, J. & Bandettini, P. A. How long to scan? The relationship between fMRI temporal signal to noise ratio and necessary scan duration. Neuroimage 34, 565–574 (2007).

59. Andersson, J. L. R. & Sotiropoulos, S. N. An integrated approach to correction for off-resonance effects and subject movement in diffusion MR imaging. Neuroimage 125, 1063–1078 (2016).

60. Fortin, J.-P. et al. Harmonization of cortical thickness measurements across scanners and sites. Neuroimage 167, 104–120 (2018).

